# Epidemiological investigation of the COVID-19 outbreak in Vellore district in South India using Geographic Information Surveillance (GIS)

**DOI:** 10.1101/2022.04.21.22274138

**Authors:** Malathi Murugesan, Padmanaban Venkatesan, Senthil Kumar, Premkumar Thangavelu, Winsley Rose, Jacob John, Marx Castro, Manivannan T, Venkata Raghava Mohan, Priscilla Rupali

## Abstract

**Objectives:** Geographical Information Surveillance (GIS) is an advanced digital technology tool that maps location-based data and helps in epidemiological modeling. We applied GIS to analyze patterns of spread and hotspots of COVID-19 cases in Vellore district in South India.

**Methods:** Laboratory-confirmed COVID-19 cases from the Vellore district and neighboring taluks from March 2020 to June 2021 were geo-coded and spatial maps were generated. Time trends exploring urban-rural burden with an age-sex distribution of cases and other variables were correlated with outcomes.

**Results:** A total of 45,401 cases of COVID-19 were detected with 20730 cases during the first wave and 24671 cases during the second wave. The overall incidence rates of COVID-19 were 462.8 and 588.6 per 100,000 populations during the first and second waves respectively. The pattern of spread revealed epicenters in densely populated urban areas with radial spread sparing rural areas in the first wave. The case fatality rate was 1.89% and 1.6% during the first and second waves that increased with advancing age.

**Conclusions:** Modern surveillance systems like GIS can accurately predict the trends and pattern of spread during future pandemics. A real-time mapping can help design risk mitigation strategies thereby preventing the spread to rural areas.

## Background

Various epidemiological mapping systems have been employed in the past to track the spatial and temporal patterns of infectious diseases like cholera, influenza, and even plague (Sarfo and Karuppannan, 2020). During the COVID-19 pandemic, integrated and innovative surveillance methods using new technologies with accurate and speed reporting of the disease have been crucial due to the dynamic and evolving nature of the infection (Ibrahim, 2020). Geospatial technology helps alleviate this by using its different methods namely geographical information systems (GIS), global positioning systems (GPS), and other satellite mediated technology systems. GIS is a technology that is used for geographic data capture based on location which helps in mapping and epidemiological modelling using computer-assisted programs (Musa et al., 2013). In 1854, John Snow has plotted the location of Cholera patients in a hand-drawn map which linked the cases to the source (broad street pump) confirming the fact that cholera was likely waterborne (Gilbert, 1958). Over the years, there has been tremendous advancement in GIS which have provided accurate information in a real-time manner.

Geographic information surveillance (GIS) system is an advanced tool for infectious diseases and could play a vital role in analyzing spatial and temporal patterns of spread of infectious diseases. GIS mainly helps identify clusters thereby linking the social and geographical factors of disease (Musa et al., 2013). The information generated by mapping will help understand risk factors, predict the susceptible population that is going to get infected in near future, and can estimate the incidence rates of the disease. In addition, patterns of spread with the identification of high-risk populations can help formulate risk mitigation strategies by creating new healthcare infrastructure and improving existing facilities. Since there were many uncertainties regarding transmission in the initial stages of the COVID-19 pandemic (World Health Organization, 2020) with the possibility of transmission from pre-asymptomatic and asymptomatic individuals (Zou et al., 2020) we felt that it would be prudent to track the epidemiology and patterns of spread in Vellore district. In this study, we present a spatial mapping of COVID-19 infected cases in Vellore district in Tamil Nadu, South India, and adjacent taluks to identify epidemic clusters or hotspots in the community, track the pattern of spread and study the demographic characteristics of study population.

## Methodology

### Study area

In this observational cohort study, mapping the burden of disease and further analyses have been performed using the earlier undivided Vellore district region. Study regions depicting different taluks, rural administrative areas, urban town panchayats, and city corporation limits are shown in Supplementary figure 1. The undivided Vellore district (12°55′13″N 79°08′00″E) spanned over 392.62 sq. km and had a population of 906,745 as per the census 2011 data by the Government of India (Vellore City Population Census 2011-2022 | Tamil Nadu, n.d.). In the year 2020, the Vellore district was divided into three districts namely Vellore, Tirupattur, and Ranipet.

### Study population

Documented laboratory-confirmed COVID-19 patients from the Vellore district for the period 28 March 2020 to 30 June 2021 were included in the study analyses.

### Data collection

The data used for the analysis in this study was obtained from the Vellore district surveillance portal, after obtaining necessary permission from the district authorities. De identified data of all the COVID-19 laboratory-confirmed positive patients were collected from the district portal with patient address, unique epidemiological identification number, symptoms, comorbidities, complications during hospitalization and outcome.

### Spatial mapping of COVID-19 cases

The documented addresses from the district portal were geocoded to the closest neighborhoods using Google Earth Pro (Earth Versions, n.d.) by trained GIS technicians at Christian Medical College (CMC), Vellore. Spatial data on geocoordinates was linked to the available attribute data of COVID-19 cases and personal identifiers were removed from the database used for final analyses. Village and taluk level vector layers of the study region including population-level data sourced from Survey of India, which is the National Survey and Mapping Organization under the Government of India were used as base layers for spatial mapping. Using the local neighborhood level spatial information on cases, all cases were mapped onto the base layer using the software ArcMap v 10.8.1 (ArcGIS Desktop quick start guide—ArcMap | Documentation, n.d.).

### Statistical analysis

Descriptive analysis was performed to study the characteristics of the study population for age, gender, and locality (urban or rural), presentation of COVID-19 symptoms, complications and survival rate was calculated as a percentage among the study population. We analyzed the age-sex distribution of COVID-19 cases along with positivity rates by age groups, gender, and area of residence (rural/urban) with 95% CI. In addition, time trends exploring urban-rural burden and other variables were correlated with outcome data and compared between the first wave (28 March 2020 to 31 March 2021) and the second wave (01 April 2021 to 30 June 2021).

### Spatial analysis

Spatial maps depicting the distribution of COVID-19 case counts by villages and urban wards in the Vellore district were generated using the geocoded addresses and layered as points on the base map to illustrate the distribution of all COVID-19 cases from March 2020 to June 2021. A total of 42,921 cases of COVID-19 were available for spatial mapping based on the completeness of documented addresses in the study area. There was a total of 938 villages, urban localities, and reserved forest areas available as individual polygons for the Vellore district. The total number of cases falling under each of these individual area polygons was estimated by spatially joining the case layer with the polygon layer and sub-block level maps depicting caseloads for the overall period and waves 1 and 2 were generated using ArcMap. Densities of COVID-19 cases around each documented case were estimated using the point density tool within the Spatial Analyst toolbox in ArcMap and quarterly heat maps were generated to demonstrate the spread of the disease for the period March 2020 to June 2021. The earlier undivided Vellore district was subdivided into eight taluks which included Arakkonam, Wallajah, Arcot, Katpadi, Vellore, Gudiyattam, Vaniyambadi and Tirupattur spanning from west to east (Revenue Divisions | Vellore District, Government of Tamil Nadu | India, n.d.). Taluk level population counts were obtained from the recently published figures on the taluk websites and were used as denominators to estimate and map incidence rates of COVID-19 disease during the two waves.

## Results

A total of 45,401 cases of COVID-19 were detected between 28 March 2020 to 31 June 2021 with 20730 cases during the first wave (28 March 2020 to 31 March 2021) and 24671 cases during the second wave (1 April 2021 to 30 June 2021). Data was available in 42921 cases for spatial mapping based on the completeness of documented addresses during both waves of which 19,021 (44.3%) and 23,900 (55.7%) had occurred during the first and second waves respectively. A higher number of cases resided in Vellore, Katpadi and Gudiyattam taluks (Supplementary Figure 2). We mapped weekly and monthly trends in the epidemiological spread of infection and were able to identify geographical clusters, hotspots and spatial-temporal trends using GIS.

Of the 938 smaller geographical areas within the study region, 188 were classified as reserved forest areas and the rest comprised villages, urban localities, town panchayat and a city corporation located in Vellore taluk. Throughout both waves, COVID-19 cases were documented from 636 of the inhabited villages and urban areas in the Vellore district. The mean (SD) number of cases in these smaller units was 67 (278) with the minimum being one case documented in one of the villages and the maximum documented caseload was 3323 in Konavattam urban ward in Vellore city. Ten urban areas had recorded over 1000 cases each; of which one, four, and five of them were Gudiyattam, Katpadi and Vellore taluks in the region. Village and urban ward-wise caseloads during both the waves and overall study period are presented in Figure 1 and a similar trend was observed with the same urban and peri-urban neighborhoods experiencing higher burden during both the waves and with relative sparing of rural areas.

**Figure 1:**
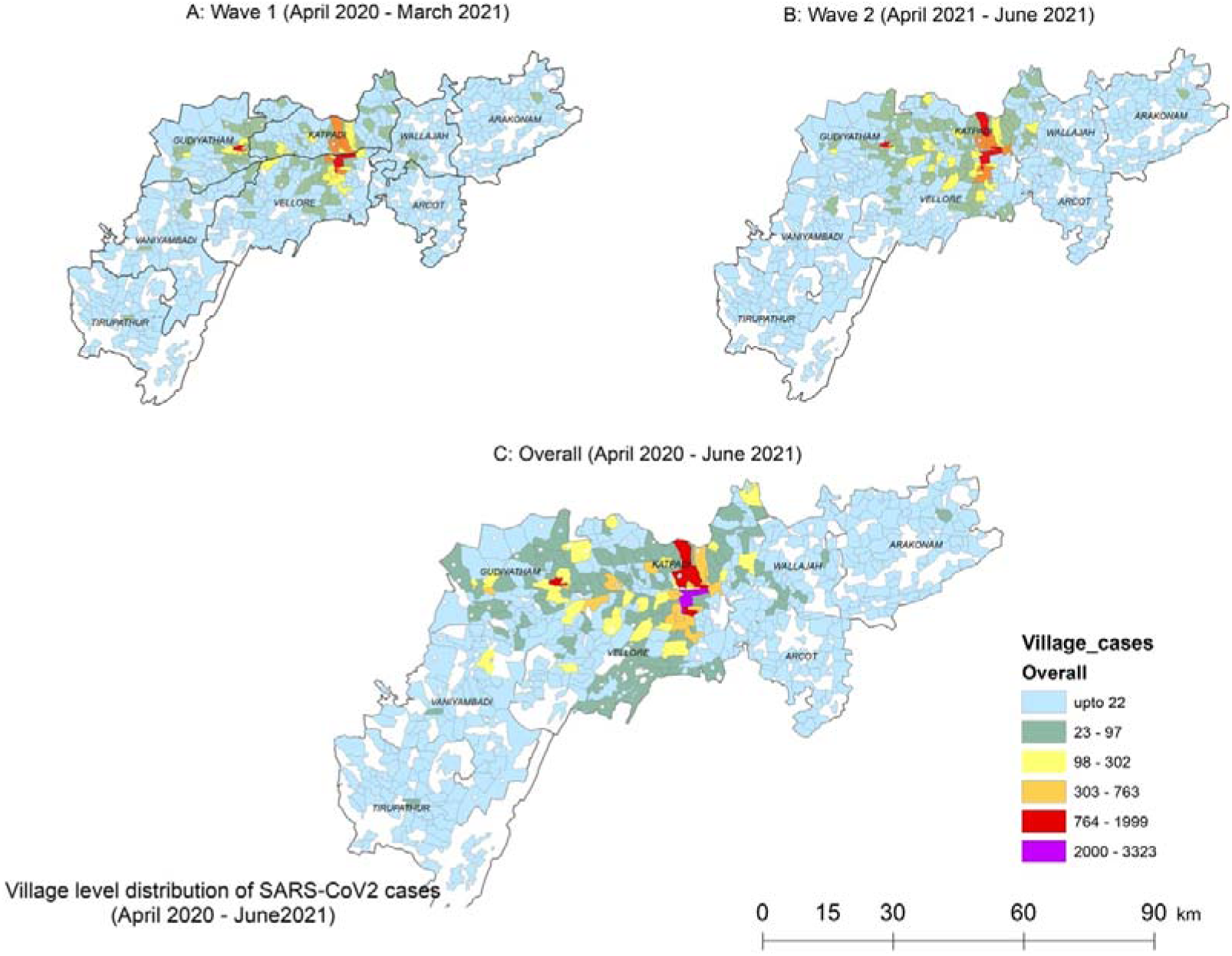
Distribution of COVID-19 cases across different geographical subunits in the study area.

Heatmaps generated estimating the point density around geolocations of cases in the initial stages of the pandemic from April to June 2020 revealed that the epicenters were in the middle of Vellore city, a densely populated city corporation and the town panchayat area of Gudiyattam taluk. Between July and September 2020 when the first wave was at its peak, it was observed that more cases occurred in the urban corporation and town panchayat areas and spread to neighboring smaller urban areas sparing the rural areas in Vellore, Gudiyattam and Katpadi taluks. Even though the other taluks (Arakkonam, Wallajah, Arcot and Tirupattur) had documented COVID-19 cases significant spatial hotspots were not detected. As the first wave started to ebb, between December 2020 and March 2021, very few neighborhoods, especially in the Vellore corporation area still had cases but at a lower density. During the second wave predominantly due to the Delta variant, which was officially documented in late March 2021, the region experienced many more COVID-19 cases as compared to the first wave. Although the pattern of spread was similar with the epicenters in urban geographies spreading radially and sparing rural areas, heat maps revealed much higher densities at these epicenters when compared to the first wave and with more peri-urban and rural area involvement (Figure 2).

**Figure 2:**
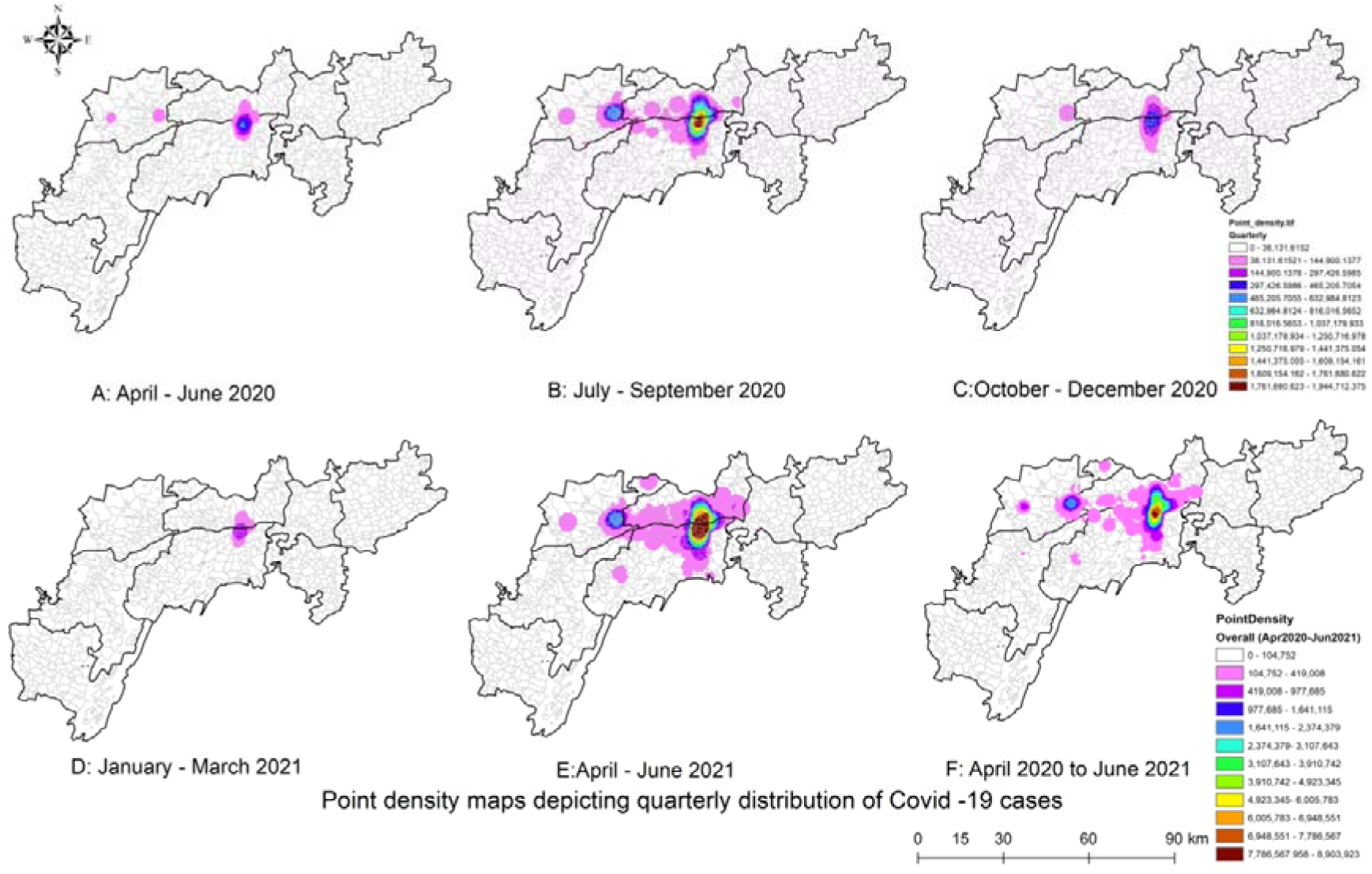
Heatmaps depicting spread of COVID-19 cases during the first and second waves in the study area.

The overall incidence rates of COVID-19 across the study region were 462.8 per 100,000 and 588.6 per 100,000 populations during the first and second waves respectively. Of the eight taluks, five of them (Tirupattur, Arakkonam, Arcot, Wallajah and Vaniyambadi) consistently had incidence rates less than 100 per 100,000 population during both the waves. During the first wave, the burden of disease in Vellore and Katpadi taluks were 1427.8 and 1344.4 per 100,000 respectively while during the second wave, the Katpadi region experienced a higher rate (2098.2 per 100,000) as compared to the Vellore region (1731.4 per 100,000 population). Incidence rates across the eight taluks during both the waves and overall study duration are presented in Figure 3.

**Figure 3:**
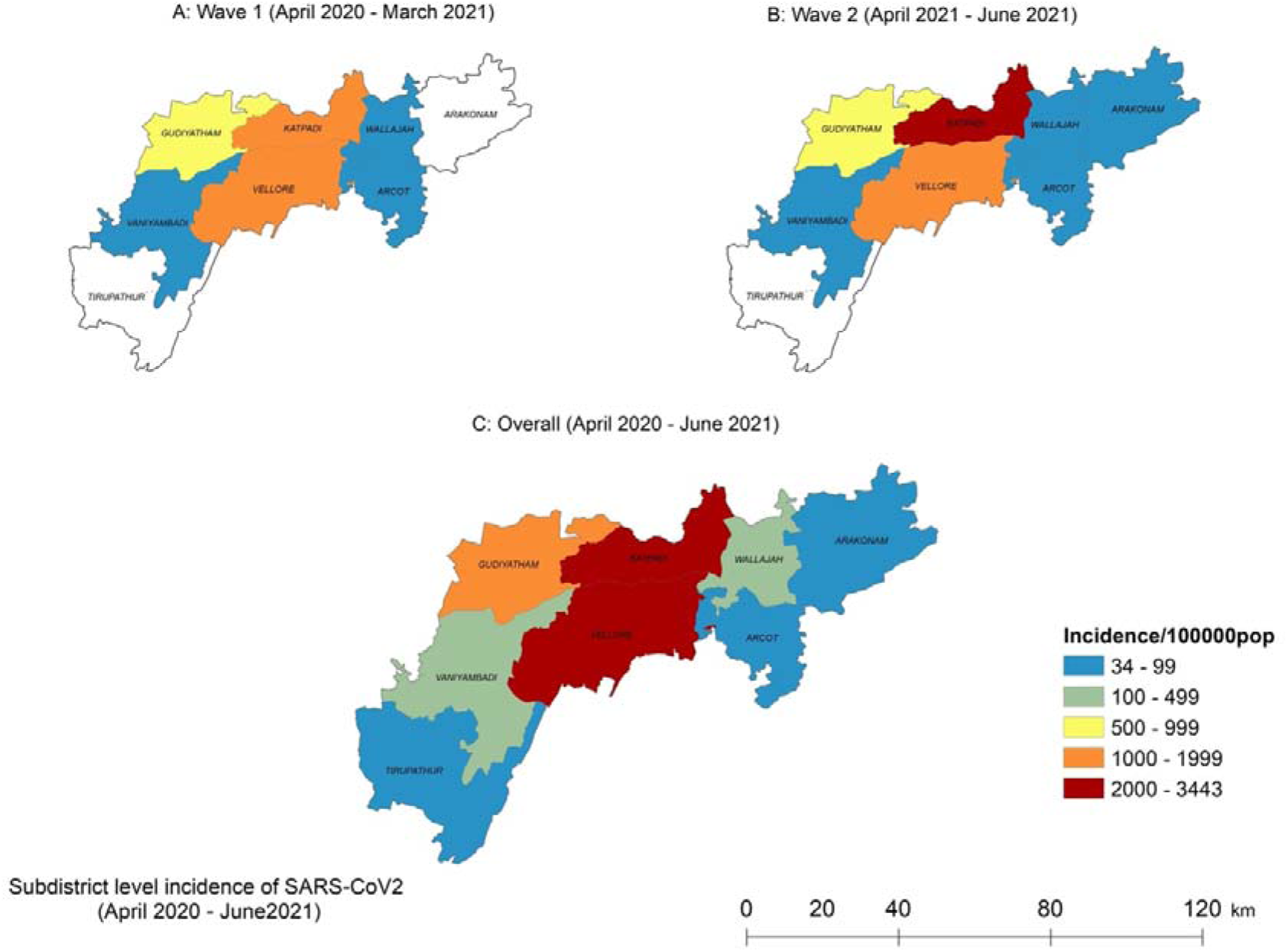
Subdistrict level incidence of COVID-19 during the first and second waves.

The age distribution among the COVID-19 cases during both the waves is given in table 1. Most of them belonged to the age group of 21-60 years. Among the study population, 3.1% in the first wave and 1.99% in the second wave were children less than 10 years. Elderly (>60 years) were 16% in the first wave and 17.3% in the second wave. Overall, COVID-19 incidence was higher in males (first wave 58.71%; second wave 56.86%) when compared to females (first wave 41.29%; second wave 42.90%). Figure 4 shows the overall time trends of the outbreak in this location. The first case documented to be COVID positive was on 28 March 2020. The outcome details were documented for 20730 cases in the first wave and 21672 cases in the second wave. Case fatality rate was 1.89% during the first wave and 1.6% during the second wave and steadily increased with advancing age, i.e., 7.38% were aged more than 60 years in the first wave and 5.02% in the second wave (Table 2). Case fatality rates were higher in men; first wave 2.40%, and 1.76% in the second wave as compared to women first wave 1.16%, second wave 1.38%. When correlating comorbidities with mortality, data from the first wave revealed that the fatality rates were highest among those who had >2 comorbidities (9.52%).

**Table 1:** Age and gender of COVID-19 patients during the first and second wave.

|  | First wave |  |  | Second wave |  |  |  |
| --- | --- | --- | --- | --- | --- | --- | --- |
| **Age**<br>**(years)** | **Female**<br>**n (%)** | **Male**<br>**n (%)** | **Total**<br>**n (%)** | **Female**<br>**n (%)** | **Male**<br>**n (%)** | **No data**<br>**n %** | **Total**<br>**n (%)** |
| 0-10 | 295 (3.4) | 349 (2.9) | 644 (3.1) | 222 (2.10) | 270 (1.92) | 0 | 492 (1.99) |
| 11-20 | 624 (7.3) | 806 (6.6) | 1430 (6.9) | 629 (5.94) | 703 (5.01) | 3 (5.00) | 1335 (5.41) |
| 21-30 | 1849 (21.5) | 2100 (17.3) | 3949 (19) | 2114<br>(19.97) | 2397<br>(17.09) | 6 (10.00) | 4517 (18.31) |
| 31-40 | 1660 (19.3) | 2297 (18.9) | 3957 (19.1) | 2128<br>(20.10) | 3115<br>(22.21) | 18 (30.00) | 5261 (21.32) |
| 41-50 | 1591 (18.5) | 2271(18.7) | 3862 (18.6) | 2017<br>(19.06) | 2543<br>(18.13) | 13 (21.67) | 4573 (18.54) |
| 51-60 | 1390 (16.2) | 2207 (18.2) | 3597 (17.4) | 1861<br>(17.58) | 2353<br>(16.78) | 10 (16.67) | 4224 (17.12) |
| 61-70 | 845 (9.8) | 1406 (11.6) | 2251 (10.9) | 1092<br>(10.32) | 1721<br>(12.27) | 8 (13.33) | 2821 (11.43) |
| 71-80 | 276 (3.2) | 570 (4.7) | 846 (4.1) | 421 (3.98) | 718<br>(5.12) | 2 (3.33) | 1141 (4.62) |
| 81-90 | 50 (0.6) | 127 (1) | 177 (0.9) | 94 (0.89) | 192 (1.37) | 0 | 286 (1.16) |
| 91-100 | 6 (0.1) | 11 (0.1) | 17 (0.1) | 7 (0.07) | 14 (0.10) | 0 | 21 (0.09) |
| Total | 8586 | 12144 | 20730 | 10585 | 14026 | 60 | 24671 |

**Table 2:**
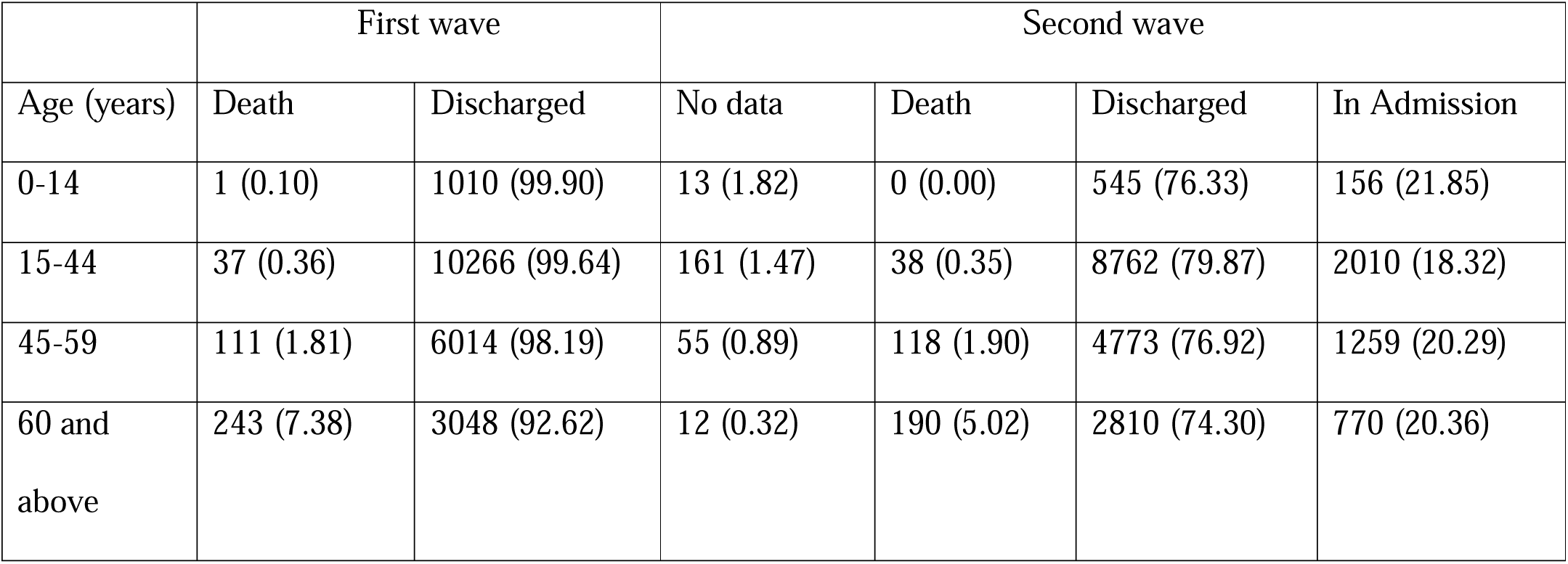
Outcome by age during first wave (N=20730) and second wave (N=21672) of COVID-19 pandemic.

|  | First wave |  | Second wave |  |  |  |
| --- | --- | --- | --- | --- | --- | --- |
| Age (years) | Death | Discharged | No data | Death | Discharged | In Admission |
| 0-14 | 1 (0.10) | 1010 (99.90) | 13 (1.82) | 0 (0.00) | 545 (76.33) | 156 (21.85) |
| 15-44 | 37 (0.36) | 10266 (99.64) | 161 (1.47) | 38 (0.35) | 8762 (79.87) | 2010 (18.32) |
| 45-59 | 111 (1.81) | 6014 (98.19) | 55 (0.89) | 118 (1.90) | 4773 (76.92) | 1259 (20.29) |
| 60 and<br>above | 243 (7.38) | 3048 (92.62) | 12 (0.32) | 190 (5.02) | 2810 (74.30) | 770 (20.36) |

**Figure 4:**
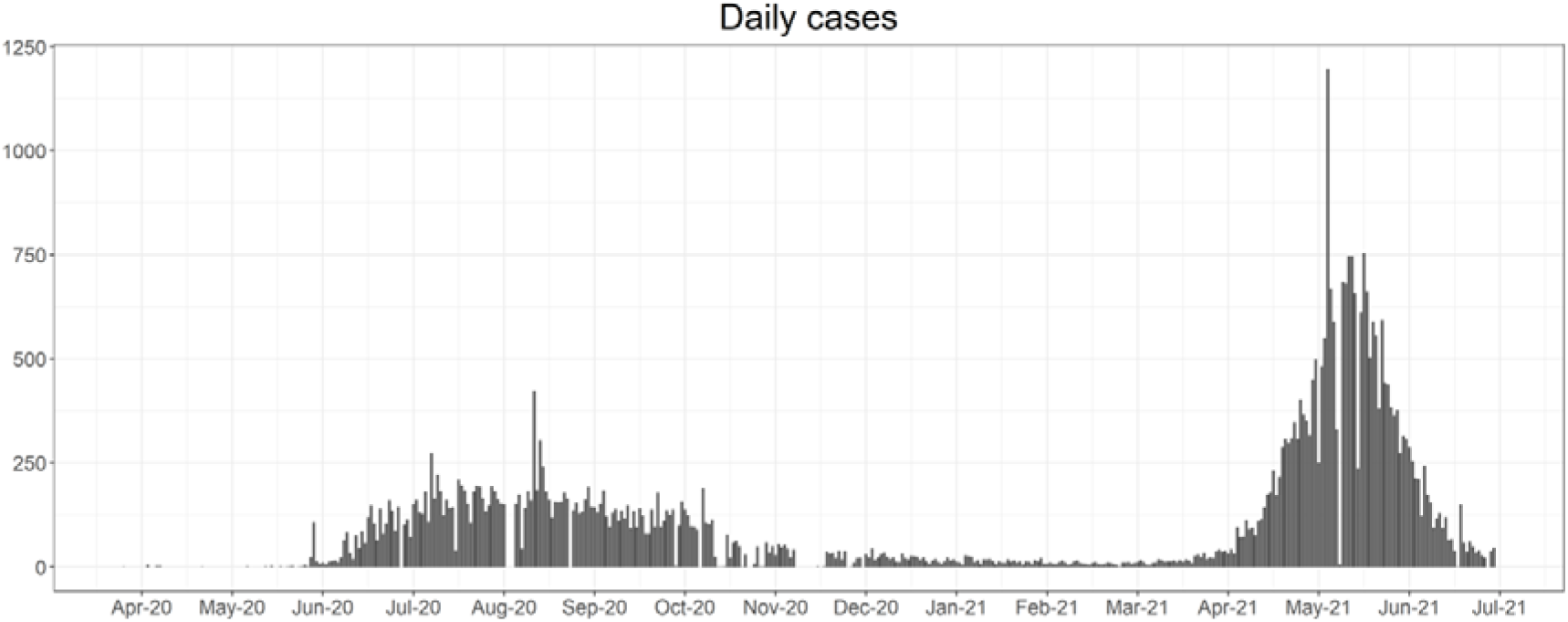
Epidemic curve of the COVID-19 pandemic during the first and the second waves.

The data on risk factors, comorbidities and clinical symptoms, and outcomes were available only for 17748 out of 19092 cases during the first wave of the pandemic but were not available for the second wave as recording this was not mandatory during the second wave. On analyzing risk factors for acquiring COVID-19 infection, 4% of them had a recent history of travel outside India and 2.1% reported a history of travel within India but 62.9% had a history of contact with a COVID-19 infected individual. The comorbidity profile data available for 17748 cases showed that 14.4% of the patients had at least one co-morbidity but 7% of the patients had more than one co-morbidity. Diabetes, hypertension, and asthma were the most seen co-morbidities in these patients (Supplementary Table 1). Among the 61.93% of patients who were symptomatic, fever (39.7%), cough (24.8%) and sore throat (15.9%) were the most common symptoms. Breathlessness was reported in 9% of all cases (Supplementary Table 2). 99 out of 17748 (0.6%) were pregnant among the study population during the first wave of the pandemic.

## Discussion

In our spatial-temporal analysis, we noted that the first wave of the pandemic started with a returned traveler from the UK in March after which a peak was noted in travelers who had attended a religious meet in another part of the country (Tablighi Jamaat, 2020). Subsequently there was a steady rise in cases over five months (June to October 2020) with continuing sporadic cases reported thereafter till March 2021. Contrary to the first wave, the epidemic noted a sharp spike in April with a rapid decline over two months (April to June 2021) during the second wave of the pandemic. This was presumed to be mainly due to the delta variant B.1.617.2 which had a high transmissibility and immune evasion as compared to the original strain (Hwang et al., 2022; Mlcochova et al., 2021). A similar pattern was observed, 100 years ago (1917-1918) when there were a successive occurrence of three waves caused by Spanish flu in one year (Taubenberger and Morens, 2006); a smaller spring wave, a larger fall wave and again a smaller winter wave. The R_0_ of Spanish flu in 1918 was 1.5 – 3.8 (Chowell et al., 2006) and the subsequent H1N1 influenza pandemic in 2009 was 1.33 (White et al., 2013). However, R_0_ for COVID-19 based on the incidence data of European countries was in fact 2.2 (95% CI 1.9-2.6) for the ancestral strain (Locatelli et al., 2021), however China reported a higher R_0_ of 3.32 (95% CI 2.81 to 3.82) (Alimohamadi et al., 2020). Subsequent waves caused by the Delta wave revealed a R_0_ of 5.08 (3.2-8) (Liu and Rocklöv, 2021) and the Omicron 9.5 (5.5-20) (Liu and Rocklöv, 2022). This suggested that R_0_ of SARS-Co-2 was second only to measles in being highly transmissible warranting strict quarantine and containment strategies for effective control. The rapid transmission left many countries and states unprepared. Hence, the usage of digital methods to map the cluster areas, predict hotspots would have enabled contact tracing in real-time helping to cordon off affected households to prevent ongoing transmission.

The geospatial mapping in our study population showed that the pattern of spread of the cases is mainly in highly populated urban areas followed by less populated semi-urban areas and then rural areas. In both the first and the second wave, the initial cluster of cases is seen in the urban population. But in the second wave, the peri-urban and rural areas were also affected. Since 60% of population in India is rural it is imperative to curb spread from urban to rural areas. Similar trend has been shown worldwide where the metropolitan cities and urban areas are affected earlier due to a higher population density, indoor crowding, and high usage of crowded public transport systems (Lee et al., 2020). A study conducted by Gangwar *et al*., analyzed COVID-19 cases in India using geospatial technology from statistics available in the public domain and showed similar results suggesting that 80% of the confirmed cases during the first wave occurred in high population areas as cities underwent gradual unlocking after a stringent lockdown phase (Gangwar and Ray, 2021).

The case fatality rate in our study population was 1.89% and 1.6% during the first wave and second wave respectively. A study published by Shah *et al*., found that the CFR for SAARC countries were lower than the developed countries and the estimated crude and adjusted CFR for India was 1.542% and 1.601% (95% CI 1.548 - 1.684) during the first wave (Shah et al., 2021). Another study by Bogam *et al*., estimated that the fatality rate was 47% lower in the delta variant driven second wave than in the first wave (adjusted CFR 0.43; 95% CI 0.41 – 0.45) (Bogam et al., 2021) despite the large numbers. This suggested that although, the transmission was higher during the second wave, the CFR is probably lower due to a predominantly younger population being affected, pre-existing immunity obtained from either previous COVID-19 infection or other coronaviruses in circulation, COVID-19 vaccination or easy access to health care services as all hospitals were directed to accept COVID-19 infected patients under the epidemic act of 1897 (The epidemic disease act, 2020, n.d.). Even though the country managed well during the first wave of the pandemic, the rapid hit of the second wave decimated the health care system (National Health Profile, 2019).

The COVID-19 pandemic has also caused an unprecedented global economic crisis due to lockdown, travel restrictions, border closures and loss of employment. The UNICEF has estimated that 42 to 66 million children could fall into extreme poverty because of the COVID-19 crisis in addition to the already existing poor population of 386 million children (2019) (UNSDG | Policy Brief, n.d.). Thus, an accurate mapping of the diseased population which could predict the spread of future respiratory pandemics would help adapt future strategies of containment and management including testing, physical distancing, contact tracing, training, and public awareness based on the resources and capacity of each health setting. Educational awareness and targeted training of public and health care sectors can be done in a phased manner in clusters/hot spots followed by other areas. The less affected areas with susceptible populations can be targeted for vaccination by doorstep delivery of vaccines through mobile camps. Even though the Epidemic diseases act 1897 was helpful in the containment of the disease spread nationwide during the pandemic, digitalized methods and real-time mapping of the clusters were not employed and thus interventions which could have helped during the second wave could not be implemented effectively. Previous respiratory pandemics have led to enormous mortality and morbidity around the world with measles epidemic in 1960s infecting 3-4 million people in the United States leading to 500 deaths (Scary measles history has been forgotten by many - The Washington Post, n.d.) and plague in India in 1900s causing over 2-3 million deaths (Plague, n.d.). This calls for appropriate measures which can accurately predict mortality, morbidity, and the course of the outbreak during future respiratory pandemics and help with real-time mapping of cases [latitude and longitude coding] on an individual case basis thereby efficiently implementing risk mitigation strategies.

As new diseases emerge and old ones re-emerge, the routinely used surveillance methods in public health include active, passive, categorical, integrated, syndromic, case-based, sentinel and serological surveillance etc. (Ibrahim, 2020; Nsubuga et al., 2006) are helpful. However, due to rapid spread of respiratory pandemics with a high R_0_ in the past, it makes sense to employ digital surveillance technology as it can save time, manpower and resources. During the pandemic, many organizations including WHO created various geospatial dashboards like the WHO Coronavirus (COVID-19) dashboard (WHO Coronavirus (COVID-19) Dashboard, n.d.), John Hopkins University COVID-19 dashboard (Coronavirus COVID-19 (2019-nCoV), n.d.), COVID-19 India (Coronavirus in India, n.d.) and many others regionally. This was extremely useful for epidemiologists and local health policymakers to implement isolation and quarantine measures based on the geographic spread. Mobile-based digital apps like the COVID symptom study tracker (ZOE COVID Study, n.d.) and Aarogya Setu (Aarogya Setu, n.d.) helped in collecting demographic characteristics of the diseased individuals but lacked real-time mapping. In India, due to the usage of the same mobile numbers in a family, network accessibility and loss of privacy due to location access limited the usage of mobile-based apps. Based on the lessons learned during this pandemic, futuristic plans, and budget allocation to more advanced information technology systems using satellite and geomapping would help in outbreak management.

### Limitations

As the analysis is based on the data available in the public domain and conducted retrospectively, geographic location of the study population (5.46%) and outcome details (6.60%) were not available for the entire study population. The clinical features and comorbidity profile was able to be retrieved only for 85.62% of the first wave population. This limited our comparison of the risk factors and clinical features during the first wave and second wave. The data captured is from the reported cases from the district portal and not from the state wise data where there may be some patients from this district who got admitted in other places. But due to lockdown and other restrictions, we believe that almost all of them were admitted within the district.

## Conclusion

Respiratory pandemics with a high R_0_ seem to start in the urban areas and then spread to rural areas. Therefore, GIS surveillance as a contact tracing digital tool would help in rapid implementation of risk mitigation and containment strategies, thus preventing spread to rural areas where access to appropriate health care services is limited. The nation must build a catastrophe resilient public health care system with well-equipped facilities under the existing National Rural Health Mission (NRHM) to manage pandemics in urban and rural areas.

## Data Availability

The data used for the analysis in this study was obtained from the Vellore district surveillance portal, after obtaining necessary permission from the district authorities.

## Ethical Approval Statement

This study conducted jointly by Christian Medical College, Vellore, and the district health authorities which is approved by the institutional review board, Christian Medical College, Vellore located in South India (IRB no:13261 dated 26.08.2020).

## Author contributions

Malathi Murugesan - Conceptualization, Methodology, Resources, Validation, Formal analysis, Data Curation, Writing-Original draft & Editing

Padmanaban Venkatesan - Methodology, Resources, Formal analysis, Data Curation, Writing-Original draft & Editing

Senthil Kumar - Methodology, Resources, Validation, Data Curation

Premkumar Thangavelu - Methodology, Resources, Validation, Data Curation

Winsley Rose - Validation, Formal analysis, Data Curation, Writing - Review & Editing

Jacob John - Validation, Formal analysis, Data Curation, Writing - Review & Editing

Venkata Raghava Mohan - Conceptualization, Methodology, Resources, Validation, Formal analysis, Data Curation, Writing - Review & Editing

Priscilla Rupali - Conceptualization, Methodology, Resources, Validation, Formal analysis, Data Curation, Writing - Review & Editing

## Acknowledgement

We wish to thank Mr. Varunkumar, Department of Community Health who assisted in geocoding and mapping.

## Supplementary tables

**Supplementary table 1:** Co-morbidities among the COVID-19 affected patients in the first wave (N = 17748)

| **Comorbidities** | **Count**<br>**(n)** | **Percentage (%)** |
| --- | --- | --- |
| Diabetes | 1427 | 8.0 |
| Hypertension | 1258 | 7.1 |
| Asthma | 848 | 4.8 |
| Liver Disease | 330 | 1.9 |
| Malignancy | 119 | 0.7 |
| Pregnancy | 99 | 0.6 |
| Chronic Renal Disease | 92 | 0.5 |
| Heart disease | 90 | 0.5 |
| Psychiatric disorders | 74 | 0.4 |
| Hypothyroidism | 72 | 0.4 |
| COPD | 57 | 0.3 |
| Post-partum (<6 weeks) | 12 | 0.1 |
| Chronic neurological or neuromuscular disease | 11 | 0.1 |
| AKI | 10 | 0.1 |
| Immunocompromised condition including HIV | 9 | 0.1 |
| Immunocompromised condition including TB | 1 | 0.0 |
| None | 15198 | 86.7 |

**Supplementary table 2:** Clinical features of COVID-19 infection in the first wave (N = 17748)

| **Clinical features** | **Count (n)** | **Percentage (%)** |
| --- | --- | --- |
| Asymptomatic | 6757 | 38.07 |
| Symptomatic | 10991 | 61.93 |
| Fever | 7042 | 39.7 |
| Cough | 4396 | 24.8 |
| Sore throat | 2828 | 15.9 |
| Breathlessness | 1615 | 9.1 |
| General Weakness | 1515 | 8.5 |
| Diarrhoea | 1468 | 8.3 |
| Headache | 637 | 3.6 |
| Nausea/Vomiting | 213 | 1.2 |
| Irritability/Confusion | 50 | 0.3 |
| Others | 3088 | 17.3 |

**Supplementary figure 1:**
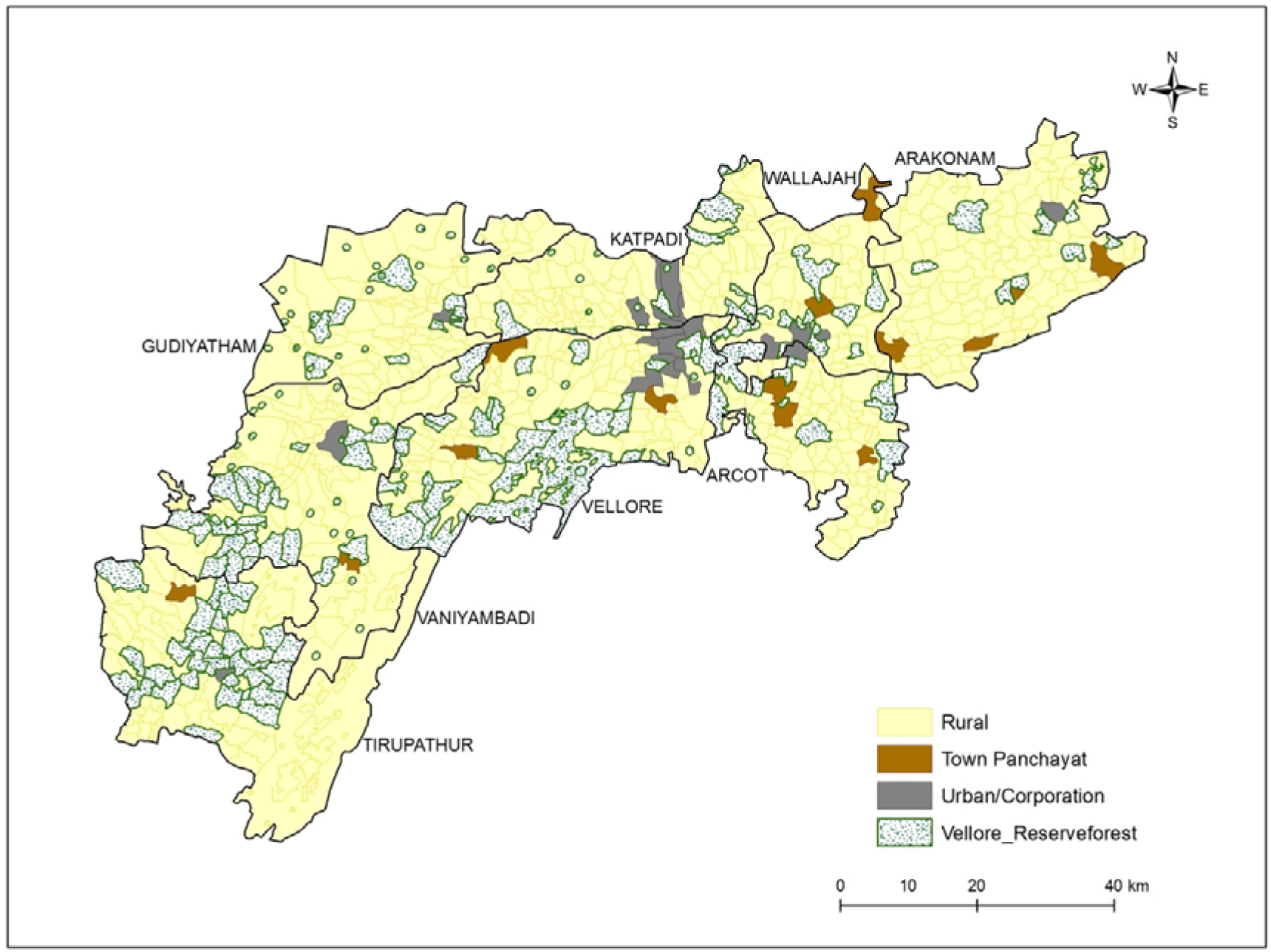
Map of undivided Vellore district showing rural and urban settlements.

**Supplementary figure 2:**
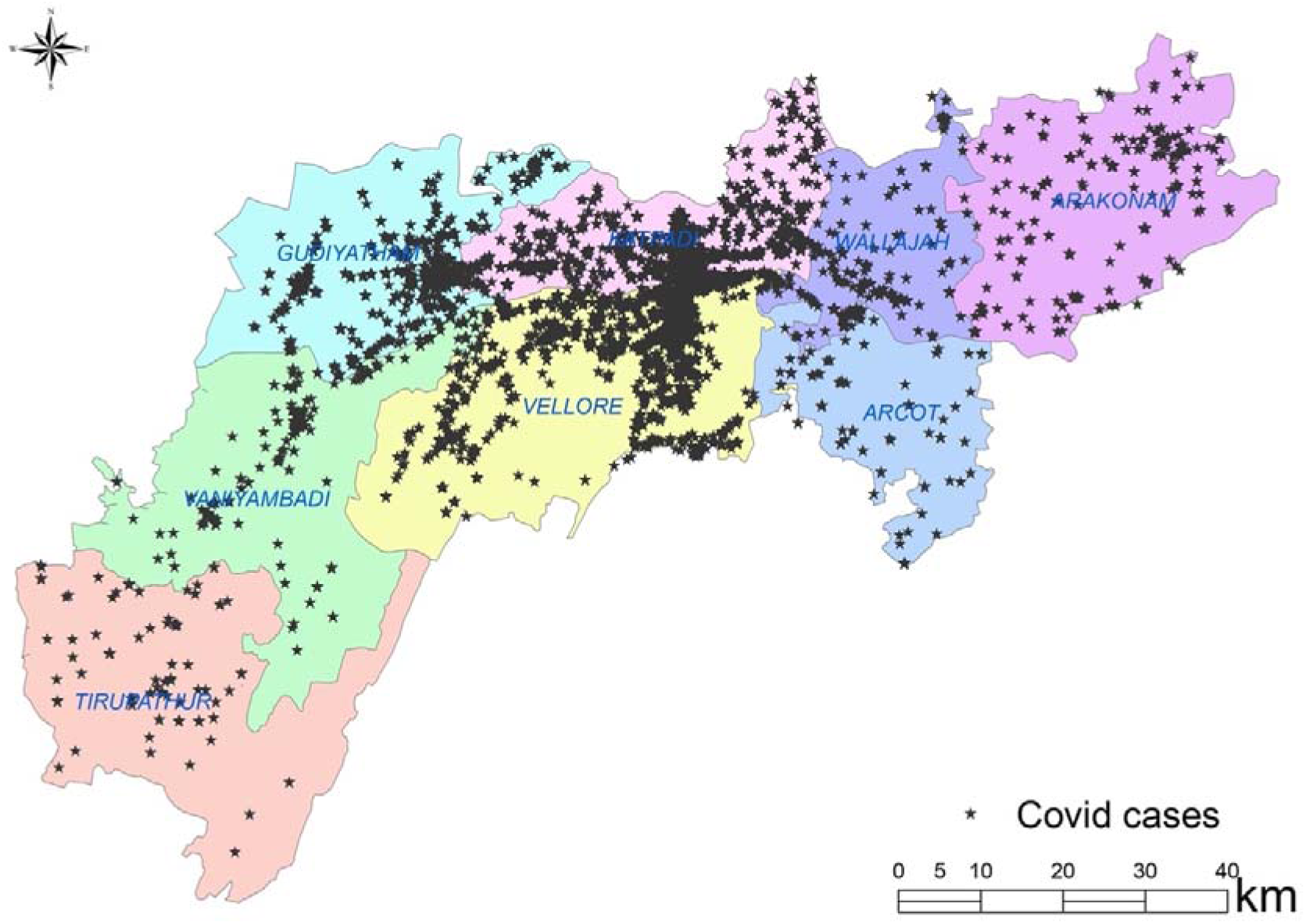
Spatial mapping of COVID-19 cases in the study region for the period March 2020 to June 2021.

